# Sex differences in the decline of neutralizing antibodies to SARS-CoV-2

**DOI:** 10.1101/2020.11.12.20230466

**Authors:** Ludivine Grzelak, Aurélie Velay, Yoann Madec, Floriane Gallais, Isabelle Staropoli, Catherine Schmidt-Mutter, Marie-Josée Wendling, Nicolas Meyer, Cyril Planchais, David Rey, Hugo Mouquet, Ludovic Glady, Yves Hansmann, Timothée Bruel, Jérome De Sèze, Arnaud Fontanet, Maria Gonzalez, Olivier Schwartz, Samira Fafi-Kremer

**Affiliations:** Virus & Immunity Unit, Department of Virology, Institut Pasteur, CNRS UMR3569, Paris, France; Sorbonne Paris Cité, Université de Paris, Paris, France; CHU de Strasbourg, Laboratoire de virologie, F-67091 Strasbourg, France; Université de Strasbourg, INSERM, IRM UMR_S 1109, Strasbourg, France; Emerging Diseases Epidemiology Unit, Department of Global Health, Institut Pasteur, Paris, France; Centre d’investigation Clinique INSERM 1434, CHU Strasbourg, France; CHU de Strasbourg, Service de santé Publique, GMRC, F-67091 Strasbourg, France; Laboratory of Humoral Immunology, Department of Immunology, Institut Pasteur, INSERM U1222, Paris, France; CHU de Strasbourg, Pôle SMO, le Trait d’Union, F-67091 Strasbourg, France; CHU de Strasbourg, Laboratoire de Biochimie Clinique et Biologie Moléculaire, F-67091 Strasbourg, France; CHU de Strasbourg, Service des infectieuses et tropicales, F-67091 Strasbourg, France; CHU de Strasbourg, Service de Neurologie, F-67091 Strasbourg, France; CHU de Strasbourg, Service de Pathologies Professionnelles, F-67091 Strasbourg, France; Vaccine Research Institute, Faculté de Médecine, INSERM U955, Université Paris-Est Créteil, Créteil, France

## Abstract

The evolution of SARS-CoV-2 humoral response in infected individuals remains poorly characterized. Here, we performed a longitudinal study of sera from 308 RT-qPCR+ individuals with mild disease, collected at two time-points, up to 6 months post-onset of symptoms (POS). We performed two anti-S and one anti-N serology assays and quantified neutralizing antibodies (NAbs). At month 1 (M1), males, individuals > 50 years of age or with a body mass index (BMI) > 25 exhibited higher levels of antibodies. Antibody levels decreased over time. At M3-6, anti-S antibodies persisted in 99% of individuals while anti-N IgG were measurable in only 59% of individuals. The decline in anti-S and NAbs was faster in males than in females, independently of age and BMI. Our results show that some serology tests are less reliable overtime and suggest that the duration of protection after SARS-CoV-2 infection or vaccination will be different in women and men.

---

The duration of humoral immune responses to SARS-CoV-2 is poorly characterized and debated. The male sex, greater age and a higher body mass index (BMI) are risk factors for a more severe disease. It is well established that severe COVID-19 patients produce higher antibody titers than asymptomatic or mildly symptomatic individuals ^1-4^. However, some studies reported stable antibody levels within the first three months of recovery, whereas others showed a rapid decrease in convalescent patients regardless of disease severity ^1-3,5-8,9,10^. Cross-sectional analyses of 10,000-30,000 individuals showed a relative stability of the humoral response ^7,11^. Anti–Spike (S) antibody amounts correlate with virus neutralization capacity, since S is the main, if not the unique, target for neutralizing antibodies. Neutralizing antibody titers also vary depending on the time post-onset of symptoms (POS) and the severity of disease ^1-4 12^. Very little is known about the influence of characteristics such as sex, age, body mass index (BMI) on the longevity and efficiency of anti-SARS-CoV-2 antibodies, particularly in mildly symptomatic individuals, who represent the majority of COVID-19 cases.

We analyzed the longitudinal antibody response in a monocentric cohort of 308 RT-qPCR confirmed staff from Strasbourg University Hospitals. The inclusion criteria are indicated in supplemental Fig. 1. The cohort included 75% females, with median age of 39 years (Table 1). The participants were nurses, doctors, caregivers and administrative staff. Contact with a COVID-19 patient, within or outside of the hospital, was reported by 37% of individuals, and 290 participants (94%) had mild symptoms consistent with COVID-19 (dry cough, fever, dyspnea, anosmia or ageusia…). Sixteen (5%) participants were hospitalized for moderate disease. None progressed to severe or critical illness. The median (interquartile range (IQR)) time from onset of symptoms to PCR testing was 3 (1-5) days. All individuals were sampled twice. The first blood sampling (M1) was performed at a median (IQR) of 31 (24-38) days POS (range: 11-58) and the second one (M3-6) at a median (IQR) of 107 (92-130) days POS (range: 78-172).

**Table 1.** Characteristics of the 308 SARS-CoV-2-infected individuals. The biological (age, sex, BMI) and clinical (contact with COVID-19 cases, reported symptoms, and time POS) characteristics of the 308 RT-PCR+ individuals are indicated. The median values are indicated, as well as the interquartile range (IQR) and total range when stated.

| Characteristics | Total<br>N (%) | Females<br>N (%) | Males<br>N (%) |
| --- | --- | --- | --- |
| Nb individuals | 308 | 232 (75.3) | 76 (24.7) |
| Age (years), median (IQR) | 39 (30.0-50.7) | 40.1 (30.0-54.1) | 34.4<br>(29.6-43.7) |
| **Age group (years)** |  |  |  |
| ≤ 30 | 77 (25.0) | 57 (24.6) | 20 (26.3) |
| 31-50 | 151 (49.0) | 109 (47.0) | 42 (55.3) |
| > 50 | 80 (26.0) | 66 (28.4) | 14 (18.4) |
| BMI (kg/m<sup>2</sup>), median (IQR) | 23.9 (21.3-27.0) | 23.9 (21.1-28.1) | 23.9 (21.6-25.8) |
| **BMI group (kg/m<sup>2</sup>)** |  |  |  |
| < 25 | 195 (63.3) | 143 (61.6) | 52 (68.4) |
| ≥ 25 | 113 (36.7) | 89 (38.4) | 24 (31.6) |
| **Contact with COVID-19 case** |  |  |  |
| No | 149 (48.4) | 113 (48.7) | 36 (47.4) |
| Yes | 115 (37.3) | 85 (36.6) | 30 (39.5) |
| Missing | 44 (14.3) | 34 (14.7) | 10 (13.1) |
| **Reported symptoms** |  |  |  |
| Aguesia and/or anosmia | 226 (73.4) | 176 (75.9) | 50 (65.8) |
| Cough | 202 (65.6) | 150 (64.7) | 52 (68.4) |
| Diarrhea | 110 (35.7) | 94 (40.5) | 16 (21.1) |
| Dyspnoea | 148 (48.1) | 117 (50.4) | 31 (40.8) |
| Fatigue | 266 (86.3) | 198 (85.3) | 68 (89.5) |
| Fever or feeling of fever | 234 (76.0) | 169 (72.8) | 65 (85.5) |
| Headache | 237 (76.9) | 185 (80.0) | 52 (68.4) |
| Myalgia | 221 (71.8) | 160 (69.0) | 61 (80.3) |
| Pharyngitis or rhinitis | 171 (55.5) | 129 (55.6) | 42 (55.3) |
| Hospitalized | 16 (5.2) | 10 (4.3) | 6 (7.9) |
| Time between onset of<br>symptoms and positive RT-qPCR test result (days),<br>median (IQR; range) | 3 (1-5; 0-14) | 3 (2-5; 0-14) | 2 (1-4; 0-14) |
| Time from onset of symptoms<br>to 1<sup>st</sup> blood sample collection<br>(days), median (IQR; range) | 31 (24-38 ; 11-58) | 32 (24.8-38; 11-53) | 27.5 (24-35;<br>13-58) |
| Time from onset of symptoms<br>to 2<sup>nd</sup> blood sample collection<br>(days), median (IQR; range) | 107<br>(92-130.2 ; 78-172) | 109.5<br>(93-131; 79-172) | 102<br>(90-126.2;<br>78-164) |

Seropositivity rates in this cohort were estimated based on time-points of collection, with four different assays (Fig. 1). We used the flow cytometry based S-Flow assay to measure anti-Spike IgG, because it displays high sensitivity (>99 %) and specificity (100%), as previously determined with pre-pandemic and SARS-CoV-2 positive samples ^13,14^. All participants had anti-SARS-CoV-2-antibodies by S-Flow at M1 and only 3 participants became negative at M3-6. The second assay was a lateral flow assay (LFA, Biosynex^™^), to detect anti-Spike IgG and IgM. This test was slightly less sensitive than the S-Flow, but stable over-time, with 85% individuals seropositive for IgG at the two time-points. IgM detection was higher at M1 (93% of seroprevalence) and decreased to 79% at M3-6, likely reflecting the contraction of the IgM response. Measurement of anti-N IgG antibodies with an ELISA (EDI^™^) gave similar results than the LFA at M1, but only 59% of individuals remained positive at M3-6. This sharp decrease may reflect a lower abundance of anti-N-specific antibodies in mild disease, a different kinetic of the anti-N response, or a lower sensitivity of the test ^15^. It has recently been reported that differences in the sensitivity of ELISA tests, including those detecting anti-N antibodies, may be dependent on the days POS ^12, 16^.

**Fig. 1.**
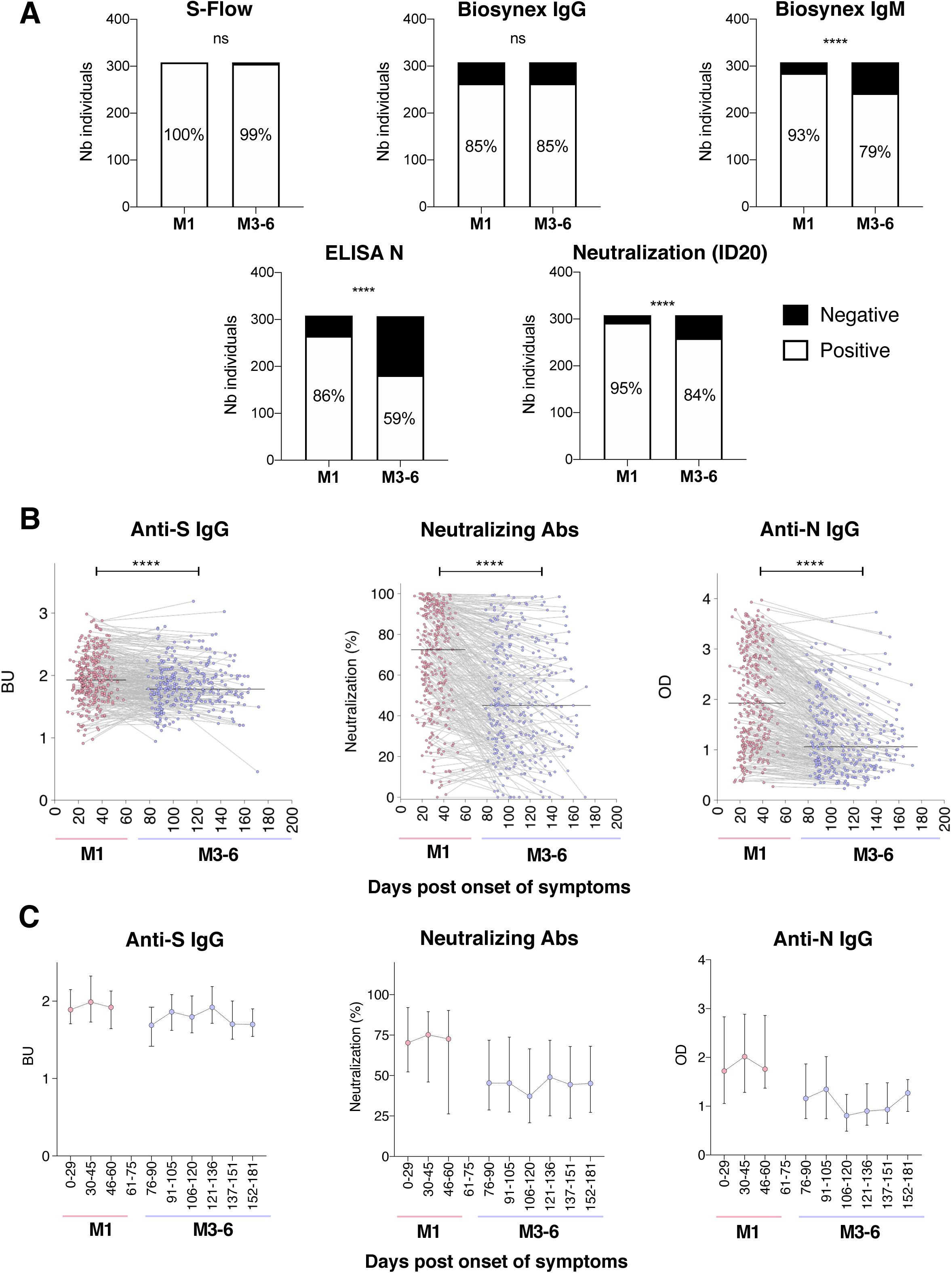
Temporal evolution of anti-SARS-CoV-2 antibodies. **A**. The number of individuals displaying anti-S (S-Flow or Biosynex™ tests), anti-N (ELISA N EDI™) or neutralizing antibodies were plotted at Month 1 (M1) and M3-6 post onset of symptoms (POS). The percentages of positive cases are indicated in the bars. Neutralization positivity was defined as a neutralizing activity against lentiviral pseudotypes higher than 20%, at a serum dilution of 1:100 (defined as Inhibitory dose (ID20)). Differences between time-points were analyzed with a Chi-square test, **** p <0.0001. **B**. The level of antibodies defined as Binding Units for anti-S IgG (S-Flow assay), percentage of neutralization, and Optical Density (OD) for anti-N IgG (ELISA) were plotted against the number of days POS. Pink and purple points stand for M1 and M3-6 time-points, respectively. Each grey line connects the two time-points from a same donor. The black line represents the median of all samples for each time-point. Paired Wilcoxon test was performed between M0 and M3-6, **** p<0.001. **C**. Donors were grouped by days POS in 14-or 29-day categories. BU, neutralization percentage and OD are shown. Results are medians with interquartile range.

We then quantified the neutralizing activity of the sera, to assess the potential protection of the humoral immune response. While different tests are currently available, we selected a neutralization assay based on lentiviral particles pseudotyped with the Spike protein. We and others previously demonstrated a strong correlation of neutralization titers obtained with single-cycle pseudovirus and infectious SARS-CoV-2 ^14,16,17^. We tested a single dilution of the sera (1:100) to calculate the neutralization activity. We previously reported a correlation between the percentage of neutralization of the lentiviral pseudotypes measured at this non saturating 1:100 dilution, and the titers obtained by performing serial dilutions of the sera ^14^. We defined a threshold of a positive neutralization at 20% (ID20 neutralization), based on background signal obtained with pre-pandemic sera (specificity of the test: 97.9%, not shown). With this threshold, 95% and 84% of the sera were positive at M1 and M3-6, respectively (Fig. 1). Applying more stringent thresholds (50% or 80% neutralization) confirmed a decline of the neutralization activity over time (Extended data Fig. 1).

We next assessed the dynamics of the immune response by comparing antibody levels at different time points POS (Fig. 2). To provide a quantitative measurement of anti-S IgG with the S-Flow, we defined a Binding Unit (BU), which corresponds to the fluorescent signal standardized by establishing a dose-response curve with a reference anti-Spike human monoclonal antibody. A longitudinal analysis demonstrated a slight but significant decrease of anti-S IgG amounts between M1 and M3-6 (Fig. 2A). This decline was also visible with neutralization activity and anti-N IgG levels of sera (Fig. 2A). We did not use the LFA in this analysis, since it does not provide quantitative results. As expected^14,16^, we observed a correlation between neutralization activity and anti-S or anti-N IgG in the sera (not shown). Plotting the median values of anti-S, neutralizing and anti-N antibodies at different time intervals confirmed this slow decline and further showed large inter-individual variations (Fig. 2B).

**Fig. 2.**
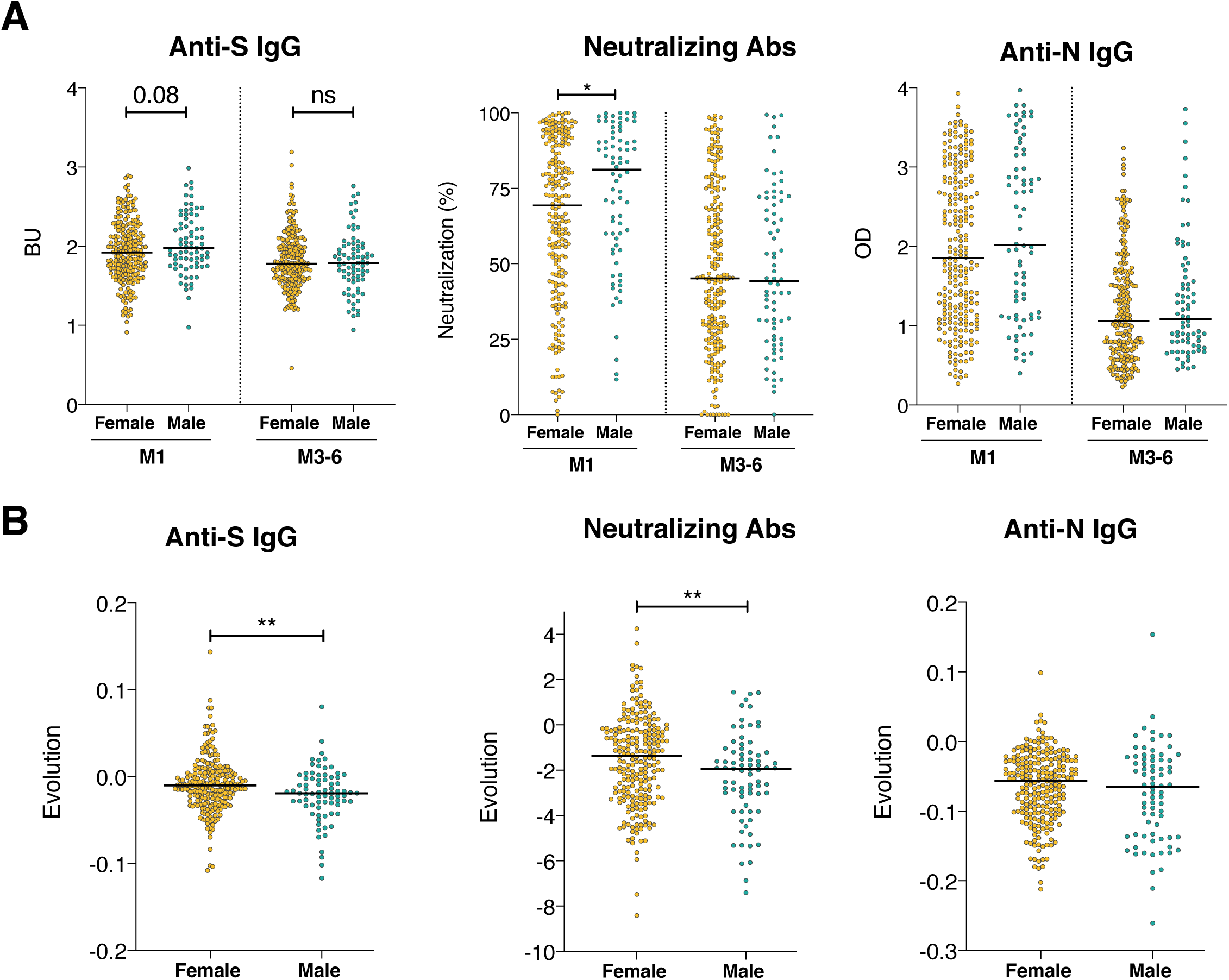
Sex differences in anti-SARS-CoV-2 antibody levels at the two samplings and their temporal evolution. **A**. Anti-S IgG (in BU), percentages of neutralization and anti-N IgG (in OD) were compared between males (green dots) and females (orange dots) at M1 or at M3-6 POS. The black line represents the median of all samples for each time-point. Samples from females and males were compared at each time-point with a Mann-Whitney test, * p<0.05, ns: not significant. p-value or non-significance (ns) are indicated on the segments. **B**. Weekly evolution of antibody levels between M1 and M3-6 was calculated as (level at M3-6 - levels at M1) / (# weeks POS M3-6 - #weeks POS M1). Color coding and graphical parameters are as in A. The dotted line represents a stable antibody level (evolution of 0). Statistical analysis Mann-Whitney test, ** p<0.01.

We then sought to determine whether variations of the antibody response may be attributed to the biological or clinical characteristics of the participants. We analyzed the associations between antibody levels (anti-S IgG, neutralizing activity or anti-N IgG) and the sex, age, BMI and type of symptoms. This analysis was performed at M1 and M3-6. We also calculated the slope of the curves between the two time-points, to assess the impact of the participants’ characteristics on the evolution of the immune response. Levels of anti-S and neutralizing antibodies were higher in males than in females at M1 (Fig. 2), in line with previous reports of a more robust induction of the immune response (cytokines and antibodies) in male patients ^18,19^. This difference was no longer visible at M3-6. Accordingly, the slope of antibody decline was significantly steeper in males (Fig. 2). A multivariate analysis showed that anti-S and neutralizing antibodies were higher at the first time-point (Extended data Table 1) and declined more rapidly in males (Table 2), independently of other parameters. There was no significant difference between males and females regarding the levels and decline of anti-N IgG.

**Table 2.** Multivariate analysis of antibody evolution. A univariate analysis of the temporal evolution of antibody levels was first performed for each of the indicated category. When the p value was < 0.15, a multivariate analysis by linear regression was performed for the indicated categories. For each type of antibodies (anti-S, anti-N, neutralizing), the factor effect with its 95% confidence interval (95% CI) and the associated p-value are indicated. Non-significant (ns) corresponds to an analysis with a univariate p-value higher than 0.15.

|  | Anti-S IgG (S-Flow) |  | Neutralizing Abs (PNT) |  | Anti-N IgG (ELISA-N) |  |
| --- | --- | --- | --- | --- | --- | --- |
|  | Factor effect (95% CI) | p-value | Factor effect (95% CI) | p-value | Factor effect (95% CI) | p-value |
| Gender |  | **0.005** |  | **0.039** |  |  |
| Female | 1 |  | 1 |  | ns |  |
| Male | -0.047 (-0.080 – -0.014) |  | -2.26 (-4.41 – -0.11) |  |  |  |
| Age (years) |  |  |  |  |  | **0.0003** |
| ≤30 | ns |  | ns |  | +0.11 (0.05 – 0.17) |  |
| 31-50 |  |  |  |  | 1 |  |
| >50 |  |  |  |  | -0.01 (-0.07 – 0.04) |  |
| BMI ≥25 |  |  |  |  |  | 0.13 |
| No | ns |  | ns |  | 1 |  |
| Yes |  |  |  |  | -0.04 (-0.09 – 0.01) |  |
| Hospitalisation |  | 0.37 |  |  |  | 0.45 |
| No | 1 |  | ns |  | 1 |  |
| Yes | -0.030 (-0.095 – 0.036) |  |  |  | -0.04 (-0.15 – 0.06) |  |
| Fever |  |  |  |  |  |  |
| No | ns |  | ns |  | ns |  |
| Yes |  |  |  |  |  |  |
| Cough |  |  |  |  |  | 0.73 |
| No | ns |  | ns |  | 1 |  |
| Yes |  |  |  |  | -0.01 (-0.06 – 0.04) |  |
| Dyspnea |  |  |  |  |  | 0.19 |
| No | ns |  | ns |  | 1 |  |
| Yes |  |  |  |  | -0.03 (-0.08 – 0.02) |  |
| Anosmia/<br>ageusia |  |  |  | 0.84 |  |  |
| No | ns |  | 1 |  | ns |  |
| Yes |  |  | -0.22 (-2.39 – 1.94) |  |  |  |
| Levels at M1 |  | **<0.0001** |  | **<0.0001** |  | **<0.0001** |
| ≤median | 1 |  | 1 |  | 1 |  |
| >median | -0.094 (-0.123 – -0.065) |  | -4.20 (-6.06 – -2.34) |  | -0.26 (-0.33 – -0.20) |  |

The majority of individuals showed a decline whereas other displayed stable antibody amounts. We categorized the subjects, based on the stability of the humoral response, into “sustainer” and “decayer” groups ^20^ (Extended data Fig. 2). The proportion of decayers varied between 71 and 83% for the three serological assays. Among decayers, the median half-life of antibody levels was 41.0 weeks (IQR: 24.3-71.8) for Anti-S IgG, 19.9 weeks (IQR: 14.4-36) for neutralizing antibodies, and 18.4 weeks (IQR: 15.2-25.7) for anti-N IgG (Extended data Fig. 2B). We also noted that female subjects were in higher proportion sustainers than decayers compared to males (Extended data Fig. 2C), in line with our observation that antibodies persist for longer periods of time in women (Fig. 2B).

Categorization of the participants by age (≤30, 30-50 and >50 years old) and by BMI (17-25, ≥25) further showed that older participants and those with a high BMI had higher antibody titers at M1, as seen with anti-S, neutralization and anti-N antibodies (extended data Fig. 3). However, the decline of antibody levels occurred at the same rate, regardless of age or BMI (Table 2 and extended data Fig. 3).

There was no association of reported clinical signs, except anosmia/ageusia, to the amount of antibodies at M1 nor with their evolution overtime (extended data Table 1 and Table 2). This likely reflects the homogeneity of symptom severity in this cohort, as all participant suffered from a mild-to-moderate disease. A minor fraction of patients were temporarily hospitalized (n=16). As previously reported ^14,21^, the antibody levels at M1 were higher in hospitalized individuals but they decreased at the same rate than non-hospitalized patients (extended data Table 1 and extended data Fig. 4), Multivariate analyses indicated that high antibody levels at M1 were associated with a more rapid decline, independently of any other parameters (Table 2). This is in line with a recently reported link between antibody persistence and disease severity ^20^.

In conclusion, we performed here a longitudinal analysis of the humoral immune response in a cohort of 308 RT-PCR confirmed SARS-CoV-2 infected patients with mild disease. We report that antibody levels and neutralizing activity decline over the 172 days of analysis. Neutralizing antibody titers decreased twice as fast as anti-S IgG, with half-lives of 19.9 and 41 weeks, respectively. Assessing the humoral response on the long-term is critical to evaluate immune protection at the population level. Commercially available assays have been validated with sera collected from acutely or recently infected individuals. We show here that some of these assays are not sensitive enough and not suited for long-term analyses. This may explain some discrepant results in the literature regarding the stability or waning of antibody titers in convalescent patients. We further report sex differences in the longevity of the immune response, with males displaying higher antibody levels shortly after infection, but a steeper decrease. Multiple studies have demonstrated that women develop more robust responses to infections and vaccination and are more sensitive to auto-immune diseases than men^22,23^. This may be linked to sex hormones, X chromosomal and environmental factors. SARS-CoV-2-infected women mount significantly more robust T cell activation than male patients^18^, which will also impact the duration of the response. Future work will help determining whether the sex differences reported here are amplified over longer periods of time, and may be linked to differences in antigen persistence ^12^. It will also be of interest extending our analysis on antibody longevity to other categories of persons, including asymptomatic individuals who represent the majority of SARS-CoV-2 cases, patients who recovered from more severe forms of COVID-19, and volunteers engaged in vaccine trials. Whether future vaccines might provide a longer protection in women than in men remains an outstanding question.

## Methods

### Study design

The objective of this longitudinal study was to assess the onset and the persistence of anti-SARS-CoV-2 antibodies in sera from mild COVID-19 healthcare workers from Strasbourg University Hospital. Among the 507 individuals of the cohort who donated samples, 308 individuals with RT-qPCR-confirmed COVID-19 diagnostic were enrolled in the study, with samples collected at two timepoints: Month 1 (M1) (median 31 days, range 11-58 days) and M3-6 (median 107 days, range 78-172 days) POS. Primary data are provided in the figures or the Supplementary Materials.

### Participants

Since April 2020, a prospective, interventional, monocentric, longitudinal, cohort clinical study enrolling hospital staff from the Strasbourg University Hospitals is on-going (ClinicalTrials.gov Identifier: NCT04441684). At enrolment (From April 15 to 29 2020) written informed consent was collected and participants completed a questionnaire which covered sociodemographic characteristics, virological findings (SARS-CoV-2 RT-PCR results including date of testing) and clinical data (date of symptom onset, type of symptoms, hospitalization). The symptoms included in the survey were: myalgia, shortness of breath or difficulty breathing, fever, asthenia, rhinitis/pharyngitis, cough, headache, anosmia/dysgeusia, diarrhea, other. Laboratory identification of SARS-CoV-2 was performed at least 10 days before inclusion by RT-PCR testing on nasopharyngeal swab specimens according to current guidelines (Institut Pasteur, Paris, France; WHO technical guidance). The assay targets two regions of the viral RNA-dependent RNA polymerase (RdRp) gene with a threshold of detection of 10 copies per reaction.

### Cells

293T cells (ATCC® CRL-3216™) were grown in complete DMEM medium (10% Fetal Calf Serum, 1% Penicillin/ streptomycin). 293T cells were transduced to express SARS-CoV-2 spike protein (GenBank: QHD43416.1) or were transduced with an empty lentivector to assess background staining. The transduced cells were selected with 2.5 ug/mL of puromycin. 293T cells stably expressing ACE2 and inducible TMPRSS2 (293T-ACE2-iTMPRSS2) were made by lentiviral transduction and selection with puromycin (1 ug/mL) and blasticidin (10 ug/mL). Cells were tested for absence of mycoplasma contamination using Mycoalert™ Mycoplasma Detection Kit (Lonza).

### Serological assays

#### Commercial serological assays

All serum samples were first tested at Strasbourg University Hospital using two commercial assays:

1) a CE-Marked LFA for detection of IgM and IgG against the SARS-CoV-2 RBD of the S protein developed by Biosynex® (COVID-19 BSS IgG/IgM). The test has a specificity of 99% and a sensitivity of 96% for samples >22 days POS ^24^. Sera were tested according to manufacturer’s instructions. Results were scored according to the sample lines intensity compared to those of the control line. The absence of the sample line was scored as negative, whereas a visible sample line was classified as positive. Interpretation was done by two independent readers.

2) a CE-Marked ELISA assay for detection of IgG against the full-length recombinant N protein from Epitope Diagnostics (EDI^™^ Novel coronavirus COVID-19 IgG). The test has a specificity of 96% and a sensitivity of 81% after 28 days POS in our hands ^24^. Cutoffs for IgG detection were calculated according to the manufacturer’s instructions.

#### S-Flow assay

A first version of the S-Flow assay was previously described ^14^ and was adapted by using 293T cells stably expressing the codon-optimised SARS-CoV-2 spike protein (293T Spike cells). Stainings were also performed on control (293T Empty) cells. The specificity and sensitivity of this assay were assessed using 253 pre-pandemic samples and 377 RT-PCR confirmed SARS-CoV-2 cases. The sensitivity is 99.2% with a 95% confidence interval of 97.69%-99.78% and the specificity is 100% (98.5-100). As previously described, 5×10^4^ cells were plated in a 96-well round bottom plate and 50 µL of a 1:300 dilution of the serum in MACS Buffer was added. The mix was incubated for 30 minutes at 4°C. The cells were then washed with PBS and stained with anti-IgG Alexa Fluor 647 secondary antibody (dilution 1:600; Thermo Fisher Scientific) for 30 min at 4°C. Cells were fixed for 10 min with 4% paraformaldehyde and data was acquired on an Attune NxT instrument (Life Technologies). Results were analysed with FlowJo 10.7.1 (Becto Dickinson & Company). After testing 253 pre-pandemic samples, the positivity of a sample was defined as a specific binding above 40%. The specific binding was calculated as follow: 100 x (% binding 293T Spike **−** % binding 293T Empty)/ (100 **−** % binding 293T Empty).

Binding units (BU) were calculated to standardize the results. A standard curve established with serial dilutions of a human anti-S monoclonal antibody (mAb 48) was acquired on the same day as the S-Flow assay. The decimal logarithm of the median of fluorescence of the sample was reported on the standard curve to obtain an equivalent value (in ng/mL) of the monoclonal antibody concentration in decimal logarithm.

### Neutralization of pseudotyped lentiviral particles

The neutralization assay was performed as described ^14^. 2×10^4^ 293T-ACE2-TMPRSS2 were plated in 96-well plates. Sera were diluted at 1: 100 and incubated with Spike-pseudotyped lentiviral particles (a kind gift from Pierre Charneau, Theravectys-Pasteur laboratory) for 15 minutes at room temperature before addition to the cells. After 48h, the luciferase signal was measured with EnSpire^®^ Multimode Plate Reader (PerkinElmer). The percentage of neutralization was calculated as follow: 100 x (1 **−** mean (luciferase signal in sample duplicate)/mean(luciferase signal in virus alone)).A titration with a human anti-S monoclonal antibody (mAb 48) was added to each plate to verify the reproducibility of the assay.

### Statistical analysis

Baseline characteristics between men and women were compared using a chi-square test for categorical variable and student’s t-test for continuous variables. Correlations between antibody measures at M1 were estimated. Factors associated with BU and neutralization levels at M1 were investigated using linear regression models, while factors associated with IgM and IgG positivity at M1 were investigated using logistic regression models. The difference in BU and neutralization levels between M1 and M3-6 was then estimated and standardized by the time interval between the two timepoints. Factors associated with these standardized differences were investigated using linear regression models. For these analyses, factors that were associated with the outcome with a p-value <0.15 in univariate analysis were introduced in the multivariate model. A p-value <0.05 was considered statistically significant. Subjects were divided into “sustainer” or “decayer” categories, for anti-S IgG, anti-N IgG and neutralizing antibody responses. To determine appurtenance at one the two categories, we calculated the fraction: antibody value at M3-6 divided by value at M1. If the fraction was ≥ 1, the donor was considered as “sustainer” for this antibody while it was considered as “decayer” if the fraction was <1. The half-life of decayers was then extrapolated from the equation of the segment formed by the two timepoints. It corresponds to the week for which the level of antibody reaches half of the M1 level.

All analyses were performed using Stata (Stat Corp., College Station, TX, USA), Excel 365 (Microsoft), RStudio Desktop 1.3.1093 (R Studio, PBC) or Prism 8 (Graphpad Software).

## Data Availability

All data are available

## Acknowledgments

We thank the patients for participation to the study, Maaran Michael Rajah for critical reading of the manuscript, the DRCI, the CIC and Médecine du travail teams for the management of the cohort, Sophie Bayer from the UCBEC team, Anne Moncolin, Veronique Sohn and Axelle Grub from the Virology laboratory for management and distribution of the samples, Pierre Charneau for the kind gift of pseudoviruses.

## Authors contribution

Conceptualization and Methodology: AF, OS, SFK

Cohort management and sample collection: AV, FG, CSM, MJW, CSM, DR, NM,YH, JDS, AF, MG, SFK

LFA and EDI tests: AV, FG, MJW, LGl

S-Flow and seroneutralization assays: TB, LG, IS, FA, PS, PC, OS

Data assembly and manuscript writing: LG, TB, YM, RG, LT, AF, SFK, OS,

Funding acquisition: AF, OS, SFK

Supervision: OS, SFK

