## Supplementary figures and table for "Sex differences in the decline of neutralizing antibodies to SARS-CoV-2"

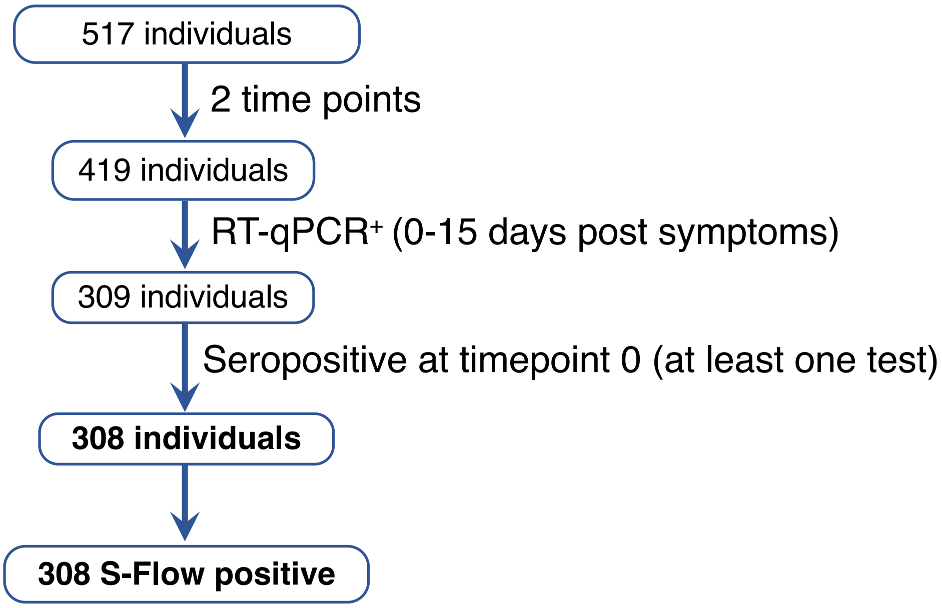


**Supplemental Fig. 1. Flow chart of the inclusion of individuals in the study.** The number of individuals appears in a blue box and the criteria of inclusion are indicated on the arrows.

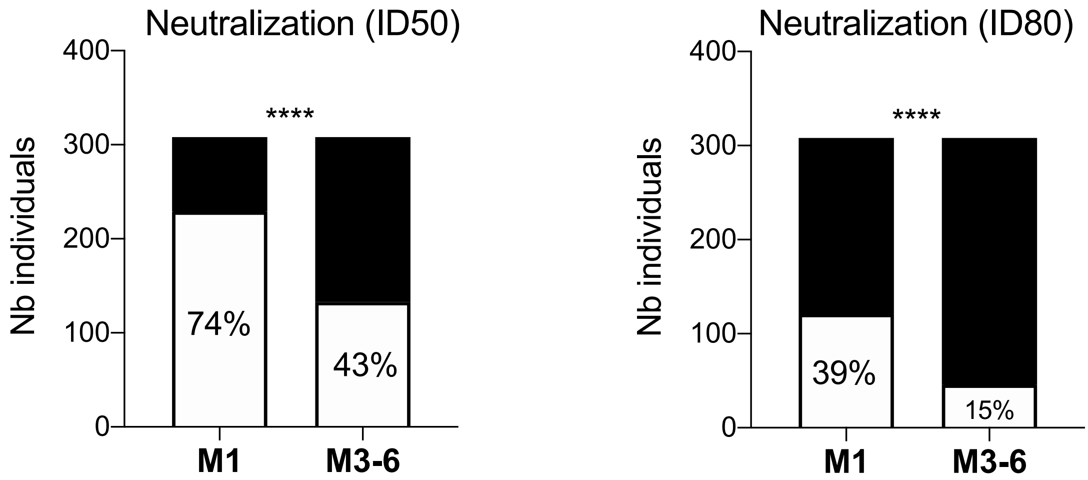


**Extended data Fig. 1. Proportion of donors with different levels of neutralizing antibodies at M1 and M3-6.** Two thresholds with different stringencies were used. The positivity was defined as a neutralizing activity against lentiviral pseudotypes higher than 50% or 80%, respectively at a serum dilution of 1:100 (defined as inhibitory dose 50 (ID50) or ID80). The number of positive and negative individuals corresponds to white and black columns, respectively. The % of positive individuals is indicated.

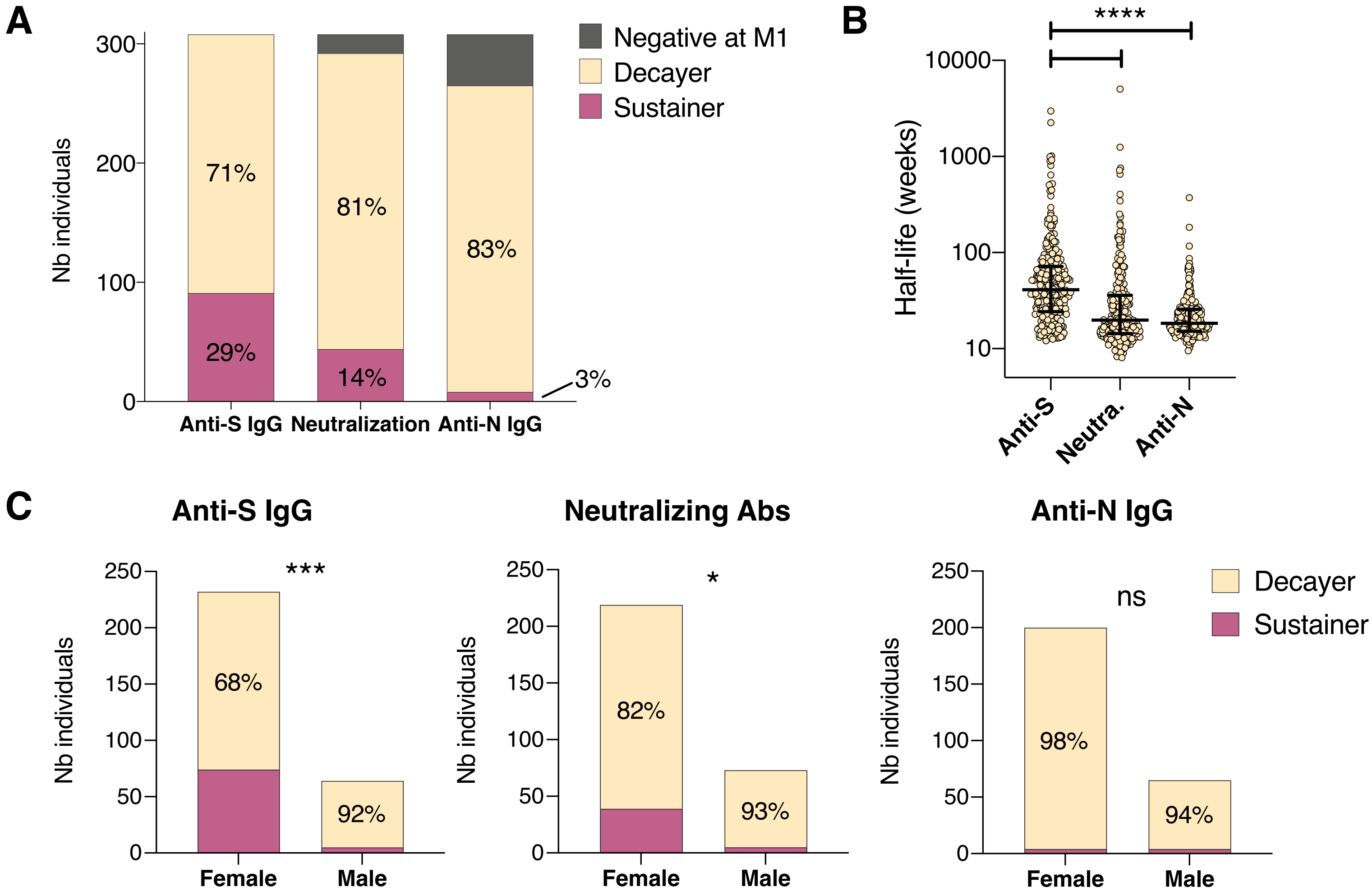


**Extended data Fig. 2. Proportion of sustainers or decayers in the cohort and half-life of antibodies among decayers. A.** For anti-S IgG, neutralizing antibodies and anti-N IgG, each subject was defined as "sustainer" (in purple) if the fraction antibody value at M3-6/ antibody value at M1≥1 or "decayer" (in yellow) if the fraction was <1. This classification was performed on donors with samples positive at M1 (donors with samples negative at M1 are in grey). The number of individuals in each category is plotted for each serological test. Statistical differences were calculated with a Kruskal-Wallis test, ****: p-value<0.0001. **B.** The antibody half-life among decayers was calculated with the equation of the segment formed by the two time-points. It corresponds to the week for which the level of antibody reaches half of the M1 level. Median and interquartile range are represented. **C.** The proportion of sustainers (in purple) and decayers (in yellow) is compared between females and males. Differences were assessed with a Chi-square test. *: p-value<0.05, ***: p-value=0.0001.

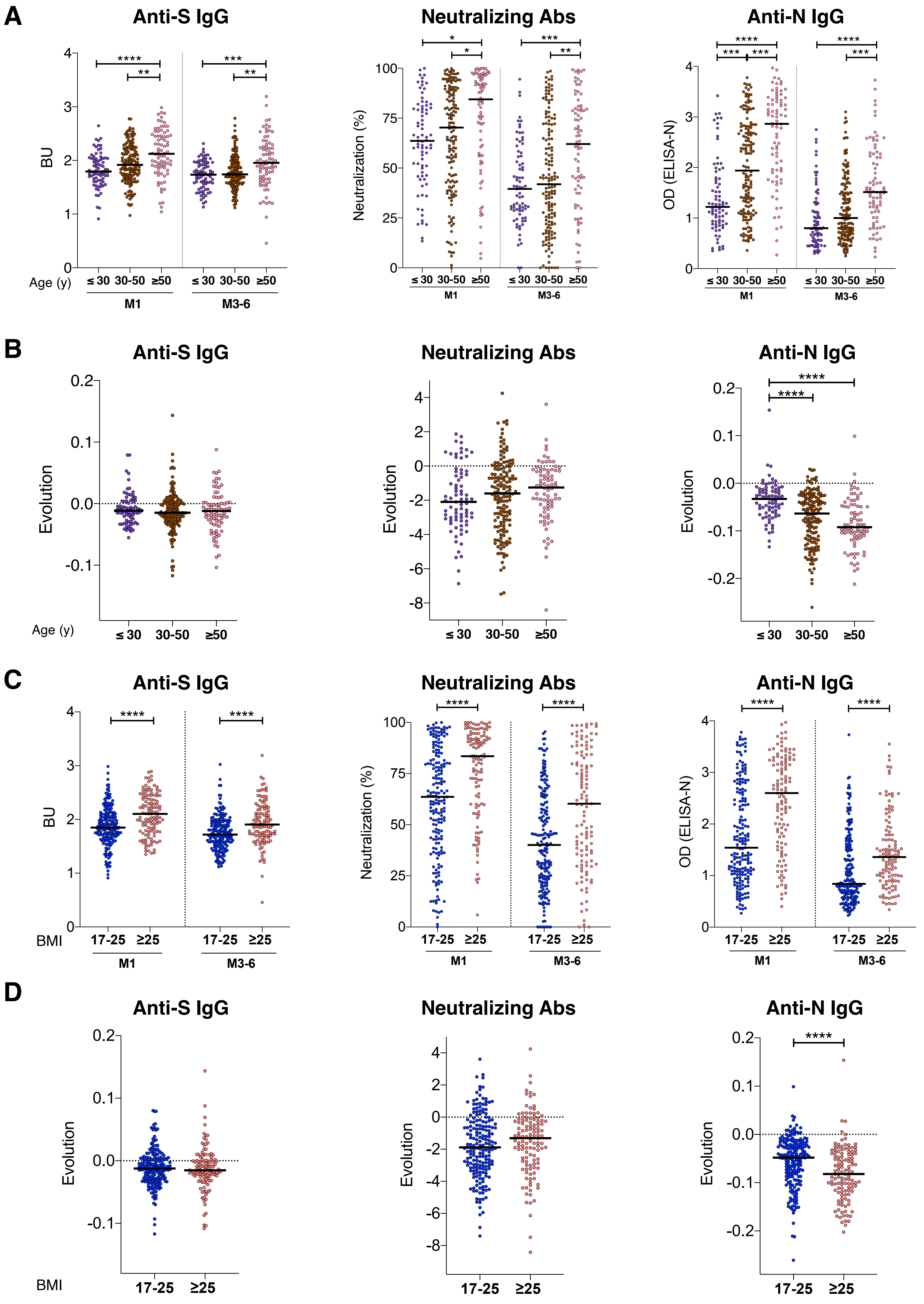


**Extended data Fig. 3. Impact of age and BMI on antibody levels and their decline with time**

**Extended data Fig. 3. Impact of age and BMI on antibody levels and their decline with time. A.** Anti S-IgG (BU), percentage of neutralization and anti-N IgG (OD) were compared between 3 age groups: <30 years old (yo) (purple dots), 30-50 yo (brown dots) and > 50 yo ((pink dots) at M1 or at M3-6 POS. The black lines depict the median value for each category. Samples at each time-point were compared using a Kruskal-Wallis test, *p<0.05, **p<0.01, ***p<0.001, ****p<0.0001. **B.** Weekly evolution of antibody levels between M1 and M3-6 was calculated for each age group as (level at M3-6 - levels at M1) / (# weeks POS M3-6 - #weeks POS M1). Color coding and graphical parameters are as in A. The dotted line represents a stable antibody level (evolution of 0). Statistical analysis Mann-Whitney test, **p<0.01. **C.** A Anti S-IgG (BU), percentage of neutralization and anti-N IgG (OD) were compared between individuals categorized in two BMI groups: 17-25 (in blue) and > 25 (in salmon). Statistical difference was assessed with a Mann-Whitney test, **** p<0.0001. **D.** Weekly evolution of antibody levels by week between M1 and M3-6 is plotted depending on the two BMI groups. Color coding and graphical parameters are as in A. The dotted line represents an evolution of 0. Statistical testing by Mann-Whitney test, **** p<0.0001.

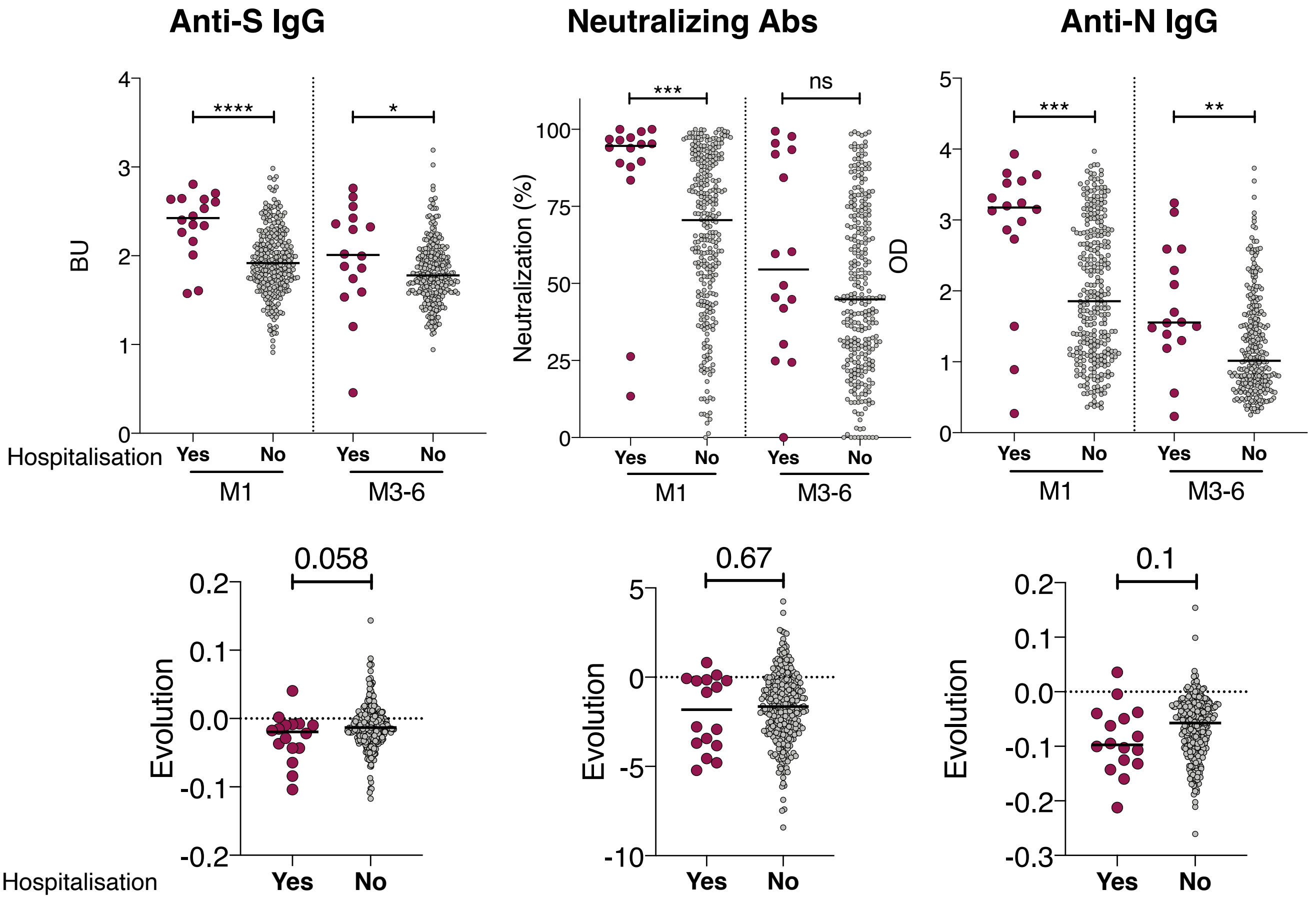


**Extended data Fig. 4. Impact of hospitalization on antibody levels and their decline with time. A.** Anti S-IgG (BU), percentage of neutralization and anti-N IgG (OD) were compared between hospitalized (red dots) and non-hospitalized (grey dots) at M1 or at M3-6 POS. The black lines depict the median value for each category. Samples at each time-point were compared using a Kruskal-Wallis test, $*p<0.05$, $**p<0.01$, $***p<0.001$. **B.** Weekly evolution of antibody levels between M1 and M3-6 was calculated for each age group as (level at M3-6 - levels at M1) / (# weeks POS M3-6 - #weeks POS M1). Color coding and graphical parameters are as in A. The dotted line represents a stable antibody level (evolution of 0). Statistical analysis: Mann-Whitney test, $**p<0.01$ ****: p-value<0.0001. When non-significant, the p values are indicated.

| | Anti-S IgG (S-Flow) | | Neutralizing Abs (PNT) | | Anti-N IgG (ELISA-N) | |
|---|---|---|---|---|---|---|
| | Factor effect (95% CI) | p-value | Factor effect (95% CI) | p-value | Factor effect (95% CI) | p-value |
| Delay since symptoms | | 0.15 | | **0.016** | | 0.81 |
| ≤28 days | 1 | | 1 | | 1 | |
| >28 days | +0.06 (-0.02-0.15) | | **-7.09 (-12.86 – -1.32)** | | -0.02 (-0.22 – 0.17) | |
| Gender | | | | | | |
| Female | 1 | **0.032** | 1 | **0.025** | 1 | **0.037** |
| Male | **+0.10 (0.01 – 0.19)** | | **+7.19 (0.92 – 13.46)** | | **+0.24 (0.01 – 0.46)** | |
| Age (years) | | | | | | |
| ≤30 | **-0.06 (-0.16 – 0.03)** | **0.001** | +0.50 (-6.16 – 7.17) | 0.055 | **-0.51 (-0.74 – -0.27)** | **<0.0001** |
| 31-50 | 1 | | 1 | | 1 | |
| >50 | **+0.15 (0.05 – 0.25)** | | +7.95 (1.30 – 14.60) | | **+0.51 (0.28 – 0.75)** | |
| BMI >25 | | | | | | |
| No | 1 | **0.0003** | 1 | **<0.0001** | 1 | **0.0001** |
| Yes | **+0.16 (0.07 – 0.24)** | | **+14.09 (8.34 – 19.84)** | | **+0.42 (0.22 – 0.63)** | |
| Hospitalisation | | | | | | |
| No | 1 | **0.002** | 1 | 0.055 | 1 | **0.036** |
| Yes | **+0.28 (0.10 – 0.47)** | | **+12.03 (-0.26 – 24.31)** | | **+0.47 (0.03 – 0.91)** | |
| Fever | | | | | | |
| No | ns | | ns | | 1 | 0.17 |
| Yes | | | | | +0.16 (-0.06 – 0.38) | |
| Cough | | | | | | |
| No | ns | | 1 | **0.048** | 1 | 0.24 |
| Yes | | | **+5.74 (0.05 – 11.43)** | | +0.12 (-0.08 – 0.32) | |
| Dyspnea | | | | | | |
| No | ns | | ns | | ns | |
| Yes | | | | | | |
| Anosmia/agueusia | | | | | | |
| No | 1 | **0.008** | 1 | **0.011** | ns | |
| Yes | **-0.12 (-0.22 – -0.03)** | | **-8.16 (-14.44 – -1.87)** | | | |
| Ct (continuous) | ns | | ns | | ns | |

**Extended data Table 1. Multivariate analysis of M1 antibody levels.** When univariate analyses of M1 antibody levels were yielding a p-value < 0.15, a multivariate analysis by linear regression was performed. For each subset of antibodies (anti-S IgG, neutralizing, anti-N IgG), the factor effect with its 95% confidence interval (95% CI) and the p-value associated are indicated. Non-significant (ns) corresponds to an analysis with a univariate p-value higher than 0.15.
